# Containing monkeypox on a college campus: a model-based analysis

**DOI:** 10.1101/2022.09.06.22279540

**Authors:** Alexandra Savinkina, Melanie Chitwood, Jiye Kwon, Virginia E. Pitzer, Gregg Gonsalves, A. David Paltiel

## Abstract

We developed a stochastic dynamic transmission model of Monkeypox Virus (hMPXV) to simulate potential spread of hMPXV on a university campus. We used the model to evaluate containment strategies, including detection and isolation of cases and vaccination or quarantine of detected case contacts. We found that detection and isolation of cases was key to containing an outbreak and was able to avert 33%-85% of secondary infections depending on detection rate. Vaccination of known contacts was a useful strategy alongside isolation and detection and could prevent 50% of additional cases. Quarantine was found to have limited benefit.

## Background

Since May 2022, there have been over 18,000 total diagnosed monkeypox virus (hMPXV) cases in the United States (1), representing every state in the country. As colleges welcome students back to campus, there is concern that close physical contact in dormitories, athletic facilities, and other congregate settings could increase the risk of sustained hMPXV transmission.

## Objective

We sought to estimate the potential impact of isolation, quarantine, and vaccination policies on the likelihood, magnitude, and duration of a hMPXV outbreak on a college campus.

## Methods and Findings

We developed a stochastic, dynamic model of hMPXV transmission on a college campus (Appendix Figure 1). The model simulated transitions between susceptible, pre-symptomatic (non-infectious), symptomatic (infectious), and recovered states. We explored the impact of varying levels of detection and isolation of cases. We also explored two additional containment scenarios, one considering quarantine of contacts of detected cases and the other considering vaccination of contacts of detected cases. We assumed the average time from infection to symptom onset was 7.6 days and time from symptom development to recovery was 21 days. We initialized the model with 10 infections in a population of 6500 students. We assumed a basic reproductive number (*R*_0_) of 1.4 and allowed for four external infections each month.

Vaccination was assumed to be 80% effective in those who were not yet infected (2) and 40% effective at preventing symptomatic (and therefore infectious) illness among those already infected (post-exposure prophylaxis). Quarantine was assumed to last 14 days on average. Any students who developed symptoms during quarantine were immediately isolated. Any students who were infected but did not show symptoms by the end of quarantine proceeded to the infected and undetected population. Quarantined students who were not infected returned to the susceptible population.

All parameters and sources are reported in Appendix Table 1. In addition, we developed a web-based, <u>interactive implementation</u> of the model to permit college decision-makers to reproduce our findings and to customize the analysis to their institutional particulars.

In the absence of any diagnosis or isolation, the model predicted a growing outbreak (Figure 1) and up to 300 cumulative cases (∼5% of the student population) within 100 days (Appendix Figure 2). Diagnosis and isolation of 20%, 50%, and 80% of cases could avert 33%, 50%, and 85% of secondary transmissions over 100 days. Isolation capacity utilization was maximized (at an average of 25 cases in isolation) when the case detection rate was 20% (Appendix Figure 3). Fewer than 15 isolation units would be required if the diagnosis rate was 80%. Isolation capacity utilization depended on the number of infected students at start of the semester (Appendix Figure 4).

**Figure 1.**
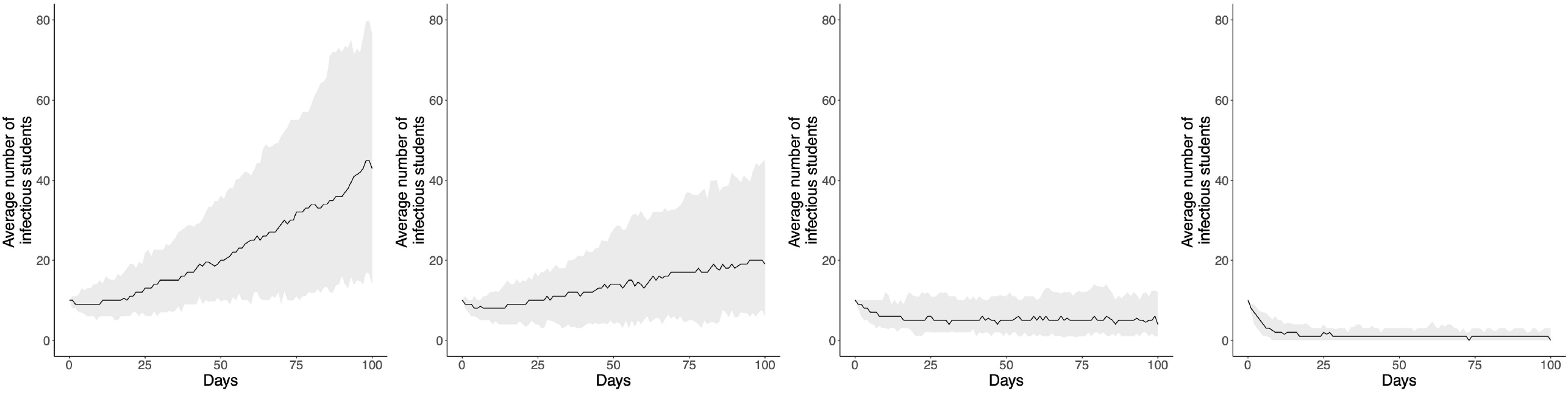
Number of monkeypox cases over time for different isolation rates. The average number of infectious students (black line) is plotted over 100 days for a population of 6,500 students with 10 infectious students at the start of semester and 4 infections from outside of the university monthly. The grey shaded regions represent the 95% prediction intervals. This scenario assumes no vaccination. Isolation varies by panel, from left to right: a. No diagnosis/isolation, b. 20% of students isolated, c. 50% of students isolated, d. 80% of students isolated.

**Figure 2.**
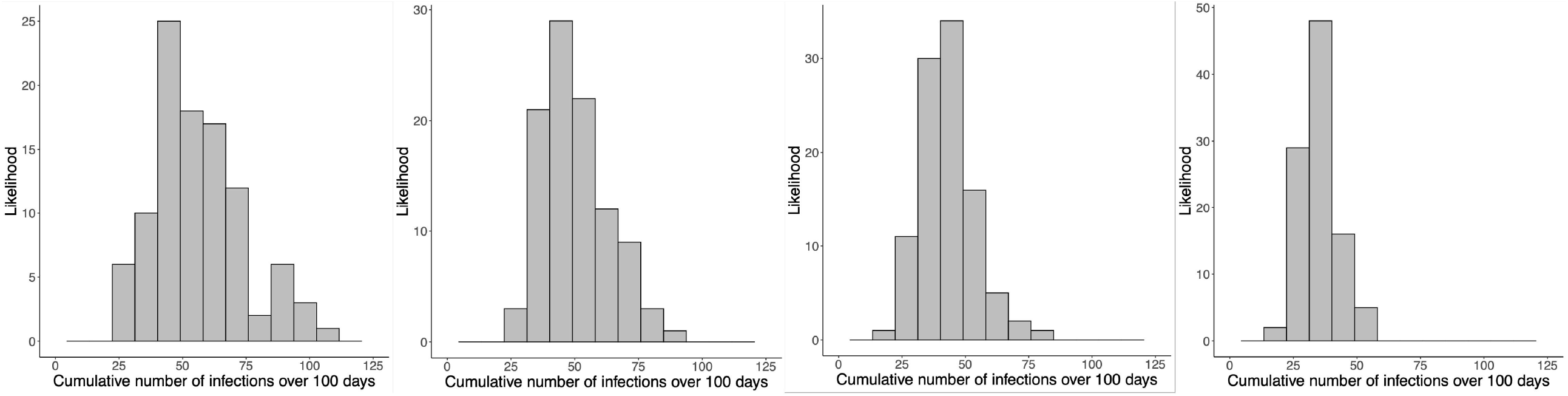
Cumulative number of monkeypox cases over 100 days for different vaccination rates. Histograms represent the distribution across 100 model simulations of the model-predicted cumulative number of student infections over 100 days in a population of 6,500 students with 10 infectious students at the start of semester and 4 infections from outside of the university monthly, with diagnosis/isolation of 50% of symptomatic cases. Vaccination varies by panel, from left to right: a. No vaccination, b. 2 students vaccinated per detected case, c. 6 students vaccinated per detected case, d. 20 students vaccinated per detected case.

Vaccination of known contacts of detected cases provided limited incremental benefit when 50% of cases were detected and isolated (Figure 2). In order to get an additional ∼50% reduction over the cases already averted by isolation, 20 contacts would need to be vaccinated per detected case (Figure 2). Up to 500 students (∼8% of the student body) would need to be vaccinated in this scenario (Appendix Figure 5).

Quarantine of known contacts provided little incremental benefit when 50% of cases were detected and isolated (Appendix Figure 6). With 20 students quarantined per detected case, as many as 200 students might need to be quarantined at one time (Appendix Figure 7).

## Discussion

Timely detection and isolation of symptomatic cases will be key to controlling hMPXV on college campuses. Since screening for hMPXV must be performed by a trained clinician – and since the estimated time from test to results is currently 3 days (3) -- educational efforts and outreach should focus on minimizing any obstacles impeding persons with symptoms from presenting for screening and care. Vaccination of known contacts may serve as a useful adjunct once vaccine supply constraints ease. We found limited benefit to quarantine of contacts, consistent with current CDC policy (4).

## Supporting information

College Monkeypox Appendix

## Data Availability

All data produced are available online at

https://github.com/ASavinkina/MonkeyPoxU

https://savinkina.shinyapps.io/MonkeyPoxUApp

