## Supplementary material for "Containing monkeypox on a college campus: a model-based analysis": College Monkeypox Appendix

**Appendix- hMPXV on University Campuses**

**Appendix Figure 1.** Model diagram.

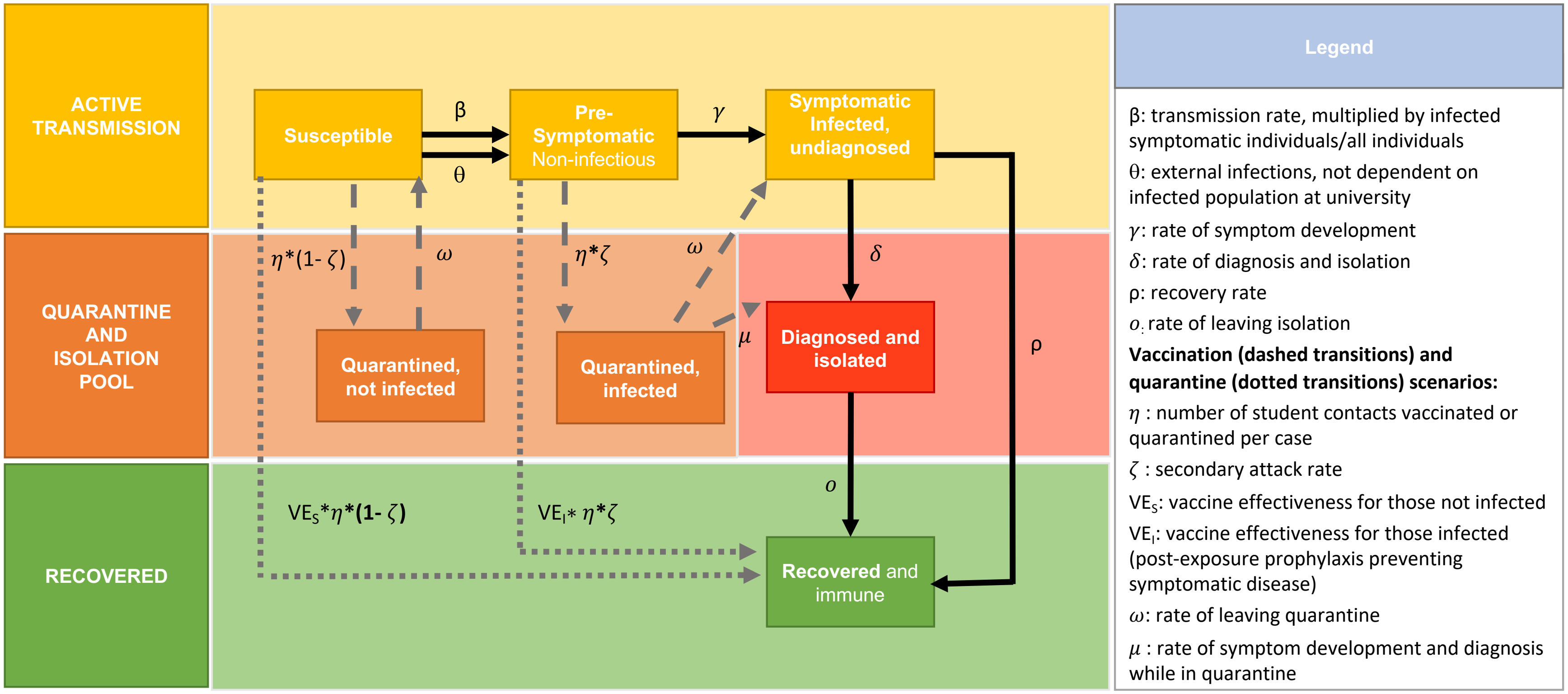

### Model differential equations:

#### With quarantine scenario:

$$\frac{dS}{dt} = -\beta(\frac{I}{N})S - \theta - \eta * (1-\zeta) * Dx_1 + \omega Qs$$

$$\frac{dP}{dt} = \beta(\frac{I}{N})S + \theta - \gamma P - \eta * \zeta * Dx_1$$

$$\frac{dI}{dt} = \gamma P + \omega Qi - \delta I - \rho I$$

$$\frac{dDx_1}{dt} = \delta I - Dx_1$$

$$\frac{dDx_2}{dt} = Dx_1 + \mu Qi - oDx_2$$

$$\frac{dQs}{dt} = \eta * (1-\zeta) * Dx_1 - \omega Qs$$

$$\frac{dQi}{dt} = \eta * \zeta * Dx_1 - \mu Qi - \omega Qi$$

$$\frac{dR}{dt} = \rho I + oDx_2$$

#### With vaccination scenario:

$$\frac{dS}{dt} = -\beta(\frac{I}{N})S - \theta - VE_S * \eta * (1-\zeta) * Dx_1$$

$$\frac{dP}{dt} = \beta(\frac{I}{N})S + \theta - \gamma P - VE_I * \eta * \zeta * Dx_1$$

$$\frac{dI}{dt} = \gamma P - \rho I - \delta I$$

$$\frac{dDx_1}{dt} = \delta I - Dx_1$$

$$\frac{dDx_2}{dt} = Dx_1 - oDx_2$$

$$\frac{dR}{dt} = \rho I + oDx_2 + VE_I(\eta * \zeta * Dx_1) + VE_S(\eta * (1-\zeta) * Dx_1)$$

**Where:**

S= Susceptible population
N = total active population (excluding isolation and quarantine)
P= Pre-symptomatic infected population, non-infectious
I = Symptomatic infectious undiagnosed population
Qs = Quarantined not infected population
Qi = Quarantined infected population
*$Dx_1$= Population newly detected and isolated (used to count number to quarantine, proceed directly to $Dx_2$ state- $Dx_1$ and $Dx_2$ together make up the detected state).
*$Dx_2$ = Infected detected and isolated population
R = Recovered and immune population

**Appendix Table 1.** Model parameter estimates and sources.

| Parameter | Estimate | In model | Source |
|---|---|---|---|
| *Total population* | 6,500 | N | Yale University undergraduate population, 2021 (1) |
| *Infected students at start of semester* | 10 | $I_0$ | Assumed |
| *Number of outside infections, monthly* | 4 | $\theta$ | Assumed |
| *Basic reproductive number ($R_0$)* | 1.4 | $\beta * 1/\rho$ | Expert opinion |
| *hMPXV secondary attack rate* | 0.2 | $\zeta$ | Estimated from $R_0$ and estimated number of contacts |
| *Time from infection to symptom onset* | 7.6 days | $1/\gamma$ | Expert opinion,<br>Miura et al.(2) |
| *Time from symptom onset to recovery* | 21 days | $1/\rho$ | Adler et al.(3) |
| *Isolation duration* | 28 days | $1/ o$ | Adler et al.(3) |
| *Quarantine duration* | 14 days | $1/\omega$ | Miura et al.(2) |
| *Vaccine Effectiveness, susceptible population* | 0.8 | $VE_1$ | Rimoin et al.(4)<br>Fine et al.(5)<br>Reynolds et al.(6) |
| *Vaccine Effectiveness, infected pre-symptomatic population (effectiveness in preventing symptomatic infection)* | 0.4 | $VE_2$ | Rimoin et al.(4)<br>Fine et al.(5)<br>Reynolds et al.(6)<br>CDC(7) |
| Percent of cases detected | 0%-80% | $\delta/(\delta + \rho)$ | Adler et al.(3) |

**Transmission model description**

We use a modified SEIR transmission model, which tracks the susceptible, infected pre-symptomatic (non-infectious), infected symptomatic (infectious), and recovered populations. At time=0, the population starts out with *N* infected individuals and total population – *N* susceptible individuals. Individuals can become infected if they are exposed to an infectious, symptomatic, undiagnosed case (*I*), at a rate $\beta*I/N$, where the transmission rate $\beta$ is equal to the estimated population-level basic reproductive number ($R_0$) divided by the average time spent in the symptomatic, undiagnosed state (=$1/(\rho+\delta)$), and *N* is the total active population (excluding those in isolation or quarantine but including those who are recovered and immune). The model also includes external introductions of hMPXV (at a rate $\theta$), allowing for students to be infected by members of the broader community.

The model includes a rate of diagnosis of infected and symptomatic students ($\delta$), which removes infected students from the infectious category. We assume that once a student is diagnosed and isolated, they will remain isolated until they have recovered and are no longer infectious, at which time they will move to the recovered and immune compartment. Students also have a chance of recovering independently from being diagnosed and isolated.

We report detection and isolation as the percent of cases diagnosed ($\delta/(\delta+\rho)$), but this can also be interpreted as the percent of time an infectious, symptomatic case spends in isolation. In other words, an 80% detection/isolation rate can be interpreted as either (a) 80% of symptomatic cases are detected and immediately isolated and the remaining 20% are never detected/isolated, or (b) 100% of cases are detected and isolated after approximately 5 days on average. The reality is likely to be somewhere in between.

We also consider the impact of vaccination on the transmission dynamics. When an infected and symptomatic case is diagnosed and isolated, students from the susceptible and pre-symptomatic populations can be vaccinated and thereby move to the immune population. The number of students removed from the susceptible vs the pre-symptomatic compartments is based on the secondary attack rate of hMPXV. Non-infected students experience a vaccine effectiveness of $VE_S$=0.8, wherein 80% of vaccinated non-infected students are fully protected from the virus and 20% experience no protection and stay in the susceptible compartment. For pre-symptomatic students, the effectiveness of the vaccine is assumed to be half the effectiveness seen in non-infected students ($VE_I$=40%) to account for students who are not vaccinated early enough in the infection (vaccination is considered most effective within 4 days of infection,(7) and the average time pre-symptomatic in the model is 7.6 days). Vaccination as post-exposure prophylaxis is modeled as preventing symptomatic (and therefore infectious) disease in 40% of vaccinated and infected individuals, and does not modify disease progression (i.e. shorten length of symptoms or decrease infectiousness and severity of symptoms) in the remaining 60% who are not protected. It is an all-or-nothing consideration – either the vaccine prevents symptomatic and infectious disease or it does not.

Finally, we model quarantine of students based on the number of diagnosed students. Case contacts move from the susceptible or pre-symptomatic compartments based on the assumed secondary attack rate of hMPXV ($\zeta$) for an average of 14 days ($1/\omega$). Those who are not infected (number of students quarantined * (1-secondary attack rate)) will return to the susceptible group. Those who are infected (number of students quarantined * secondary attack rate) will either move on to diagnosis (at a rate $\mu$ = 1/ duration of time from infection to symptom onset) or move to infected and not diagnosed (at a rate $\omega$ = 1/ duration of quarantine).

Recovered and successfully vaccinated students are considered immune and return to the general population but can no longer be infected or contribute to infection.

**Stochastic modeling**

The transmission dynamic model we developed is stochastic, allowing for variation between model runs. Stochasticity was implemented using the Gillespie stochastic simulation algorithm via the R package adaptivetau (8). This can allow us to model the likelihood of small outbreaks with $R_0$ values below 1, and also allows for modeling the probability of outbreak fade out with $R_0$ values greater than 1. While we assumed a fixed $R_0$ of 1.4, the interactive web-tool allows for $R_0$ estimates between 0.9 and 2.0. For these analyses and for the web-tool, the stochastic model is run 100 times.

**Appendix Figure 2. Cumulative number of monkeypox cases over 100 days for different isolation rates.** Histograms represent the distribution across 100 model simulations of the model-predicted cumulative number of student infections over 100 days in a population of 6,500 students with 10 infectious students at start of semester and 4 infections from outside of the university monthly. This scenario assumes no quarantine. The proportion of cases that are isolated (=$\delta/(\delta+\rho)$) varies by panel, from left to right: a. No diagnosis/isolation, b. 20% of students isolated, c. 50% of students isolated, d. 80% of students isolated.

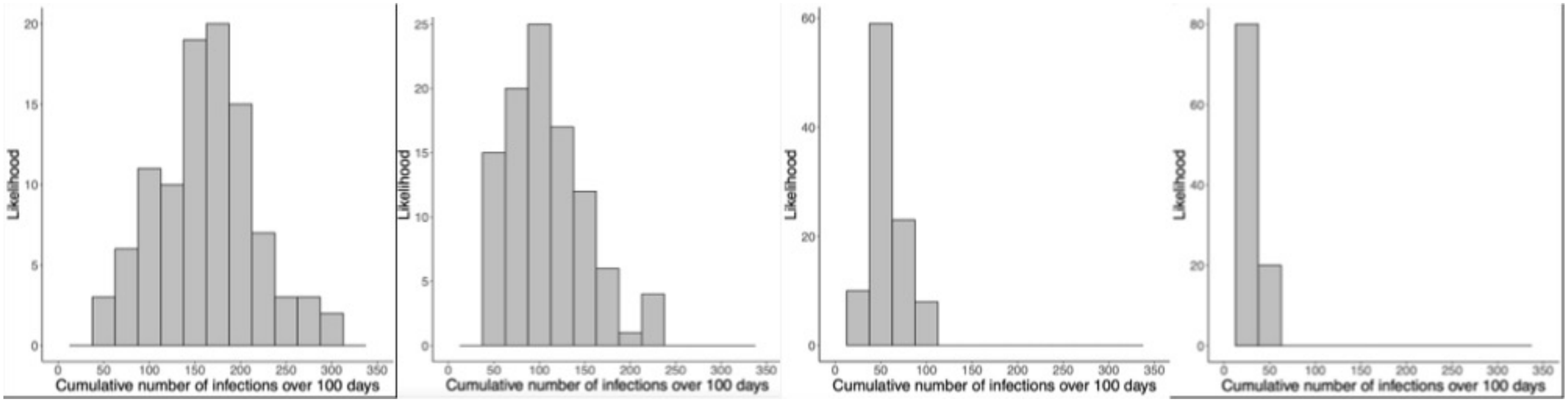

**Appendix Figure 3. Cumulative number of isolated students for different isolation rates.** Histograms represent the distribution across 100 model simulations of the model-predicted cumulative number of isolated students over 100 days in a population of 6,500 students with 10 infectious students at start of semester and 4 infections from outside of the university monthly, with no quarantine or vaccination. The proportion of cases that are isolated (=$\delta/(\delta+\rho)$) varies by panel, from left to right: a. 20% of students isolated, b. 50% of students isolated, c. 80% of students isolated.

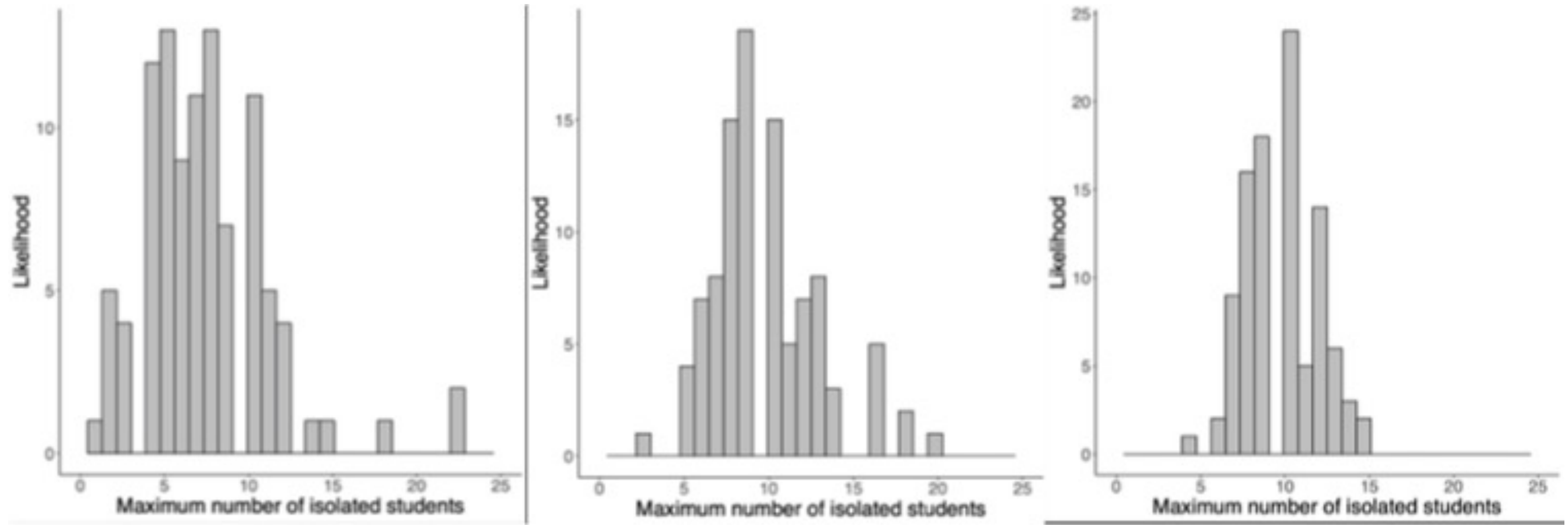

**Appendix Figure 4. Maximum number of isolated students for different values of the initial number of infections at the start of semester.** Histograms represent the distribution across 100 model simulations of the model-predicted maximum number of isolated students over 100 days in a population of 6,500 students with 4 infections from outside of the university monthly and with diagnosis/isolation of 50% of symptomatic cases. The number of initial infections varies by panel: a. 10 initial infection, b. 20 initial infections, c. 30 initial infections, d. 50 initial infections.

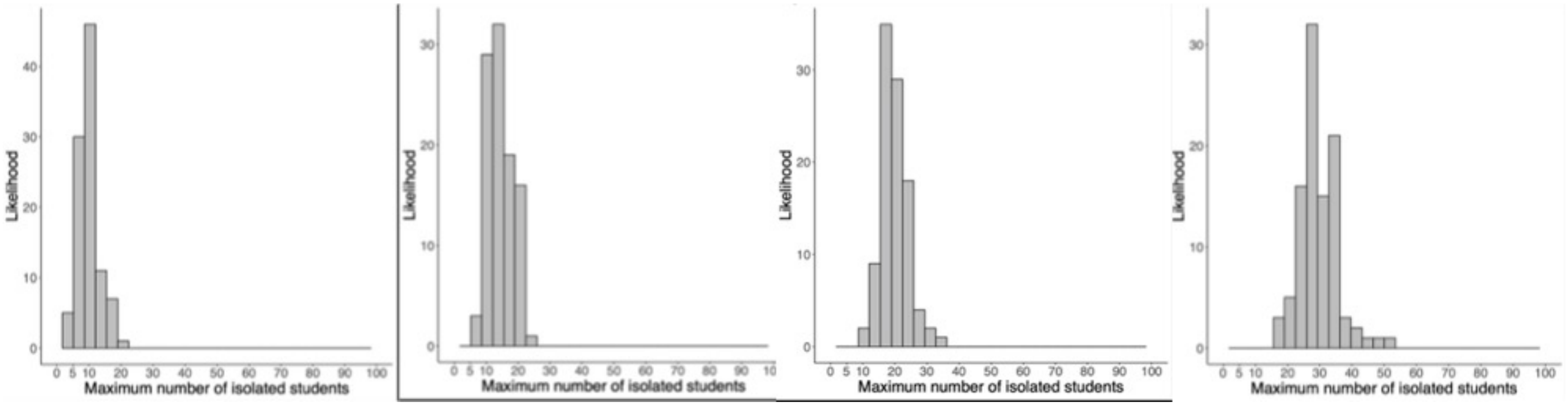

**Appendix Figure 5. Maximum number of vaccinated students for different vaccination rates.** Histograms represent the distribution across 100 model simulations of the model-predicted cumulative number of student vaccinations needed over 100 days in a population of 6,500 students with 10 infectious students at start of semester and 4 infections from outside of the university monthly, with diagnosis/isolation of 50% of symptomatic cases. Vaccination varies by panel, from left to right: a. 2 students vaccinated per diagnosed case, b. 6 students vaccinated per diagnosed case, c. 20 students vaccinated per diagnosed case.

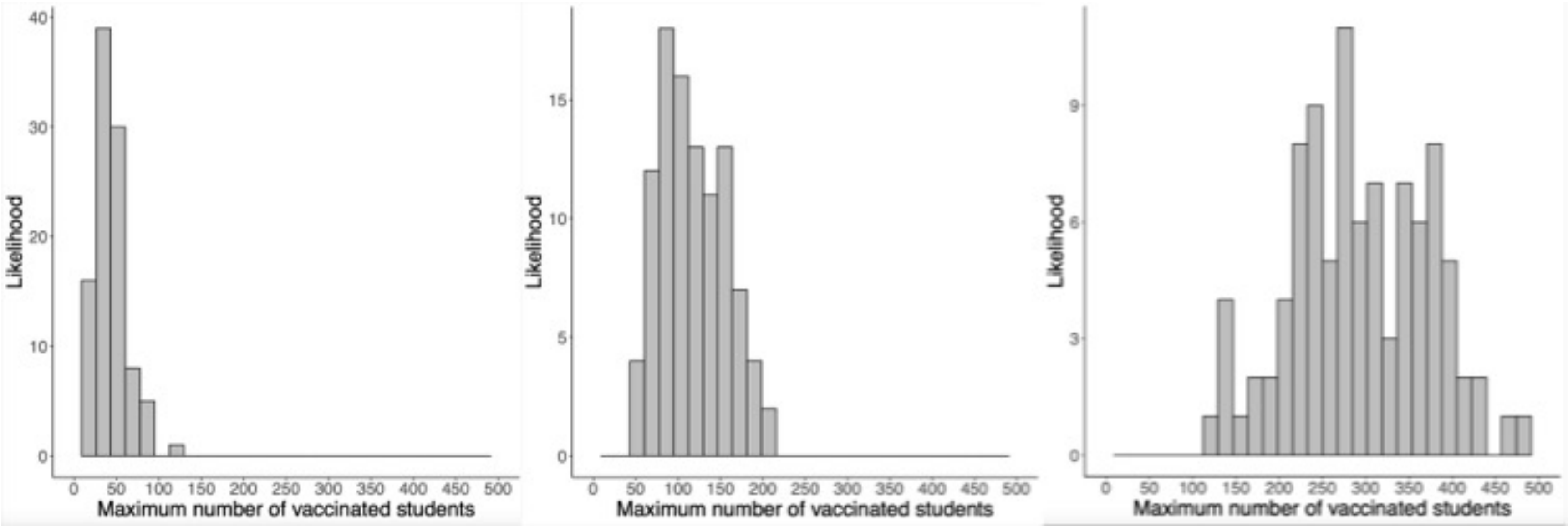

**Appendix Figure 6. Cumulative number of monkeypox cases for different quarantine rates.** Histograms represent the distribution across 100 model simulations of the model-predicted cumulative number of student infections over 100 days in a population of 6,500 students with 10 infectious students at start of semester and 4 infections from outside of the university monthly, with diagnosis/isolation of 50% of symptomatic cases and no vaccination. Quarantine varies by panel, from left to right: a. No quarantine, b. 2 students quarantined per diagnosed case, c. 6 students quarantined per diagnosed case, d. 20 students quarantined per diagnosed case.

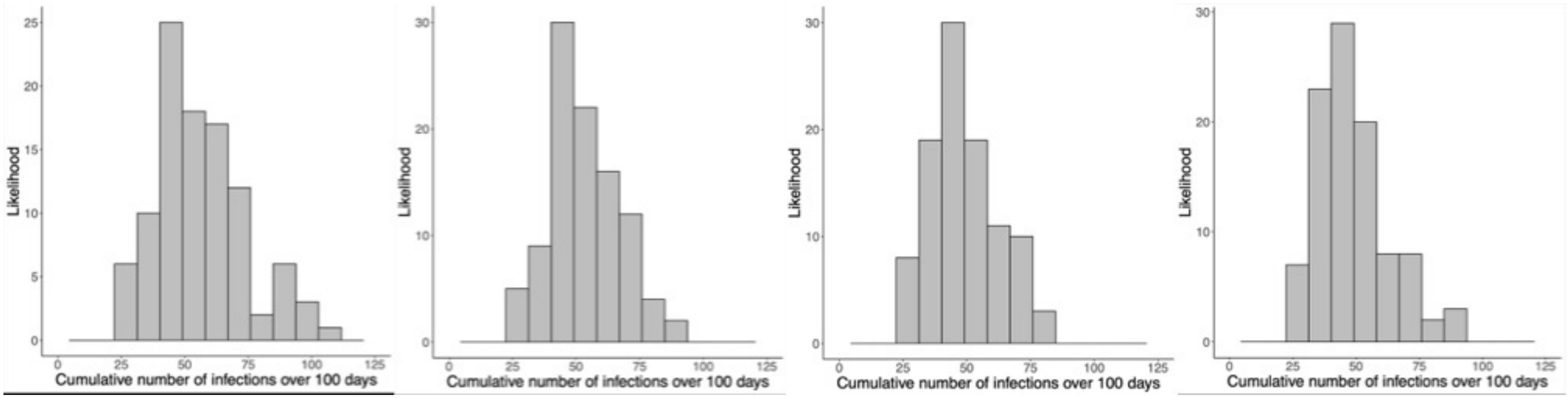

**Appendix Figure 7. Maximum number of quarantined students for different quarantine rates.** Histograms represent the distribution across 100 model simulations of the model-predicted maximum number of students in quarantine over 100 days in a population of 6,500 students with 10 infectious students at start of semester and 4 infections from outside of the university monthly, with diagnosis/isolation of 50% of symptomatic cases and no vaccination. Quarantine varies by panel, from left to right: a. 2 students quarantined per diagnosed case, b. 6 students quarantined per diagnosed case, c. 20 students quarantined per diagnosed case.

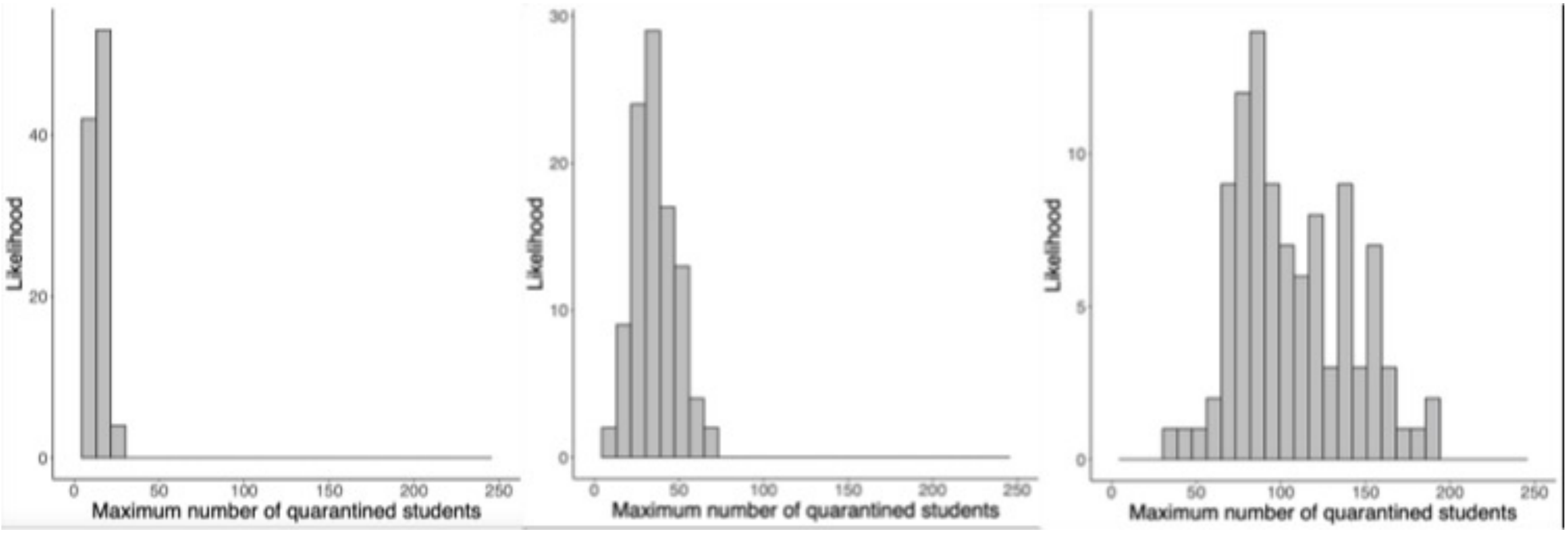
